# Combined Metabolic Activators Improves Cognitive Functions in Alzheimer’s Disease

**DOI:** 10.1101/2021.07.14.21260511

**Authors:** Burak Yulug, Ozlem Altay, Xiangyu Li, Lutfu Hanoglu, Seyda Cankaya, Simon Lam, Hong Yang, Ebru Coskun, Ezgi İdil, Rahim Nogaylar, Cemil Bayram, Ismail Bolat, Sena Öner, Özlem Özdemir Tozlu, Mehmet Enes Arslan, Ahmet Hacımuftuoglu, Serkan Yıldırım, Muhammad Arif, Saeed Shoaie, Cheng Zhang, Jens Nielsen, Hasan Turkez, Jan Borén, Mathias Uhlén, Adil Mardinoglu

**Author notes:** Lead author: Adil Mardinoglu.

## Abstract

Alzheimer’s disease (AD) is associated with metabolic abnormalities linked to critical elements of neurodegeneration. Here, we analysed the brain transcriptomics data of more than 600 AD patients using genome-scale metabolic models and provided supporting evidence of mitochondrial dysfunction related to the pathophysiologic mechanisms of AD progression. Subsequently, we investigated, in a rat model of AD, the oral administration of Combined Metabolic Activators (CMAs), consisting of NAD+ and glutathione precursors, to explore the effect for improvement of biological functions in AD. CMAs includes L-serine, nicotinamide riboside, N-acetyl-L-cysteine, and L-carnitine tartrate, salt form of L-carnitine. The study revealed that supplementation of the CMAs improved the AD-associated histological parameters in the animals. Finally, we designed a randomized, double-blinded, placebo-controlled human phase 2 clinical trial and showed that the administration of CMAs improves cognitive functions in AD patients. As decreased AD Assessment Scale-cognitive subscale (ADAS-Cog) score is the indicator of the improved cognitive function in AD patients, we observed a significant decrease of ADAS-Cog scores on Day 84 vs Day 0 (Log2FC= -0.37, (29% improvement), p-value=0.00001) in the CMA group. We also observed a significant decrease in the placebo group on Day 84 vs Day 0 (Log2FC= -0.19, (14% improvement), p-value=0.001) due to the recommendations of exercise and Mediterranean diet to all AD patients participated in the trial. A comprehensive analysis of the human plasma metabolome and proteome revealed that plasma levels of proteins and metabolites associated with redox metabolism are significantly improved after treatment. In conclusion, our results show that treating AD patients with CMAs leads to enhanced cognitive functions, suggesting a role for such a therapeutic regime in treating AD and other neurodegenerative diseases.

**HIGHLIGHTS:**

• Brain transcriptomics data of more than 600 AD patients is analysed.
• Performed an *in vivo* study using Combined Metabolic Activators (CMAs) in AD rat models.
• We performed a randomized, double-blinded, placebo-controlled human phase 2 clinical trial.
• We showed that cognitive functions in AD patients is improved 29% in the CMA group whereas 14% in the placebo group.

## INTRODUCTION

Alzheimer’s disease (AD) is characterized by progressive synaptic and axonal dysfunction, neuronal loss and cognitive decline (*1*). There is growing evidence that AD is closely associated with metabolic and oxidative stress linked to critical elements of neurodegeneration, such as mitochondrial dysfunctions and bioenergetic impairments (*2, 3*). Indeed, increasing data indicate that systemic metabolic disorders, such as insulin resistance, are strongly associated with bioenergetic failure of nerve cells(*4, 5*). This can manifest as cognitive impairment and brain-specific neuropathology while sharing common pathogenic mechanisms with AD, such as impaired glucose metabolism, increased oxidative stress, insulin resistance, and amyloidogenesis (*4, 6, 7*). Recent evidence accordingly suggests that patients with type 2 diabetes mellitus are at increased risk of developing AD (*6*).

Although the disease is still defined by the accumulation of abnormal amyloid and tau proteins (*8*), the mechanistic assumption of linear causality between the amyloid cascade and cognitive dysfunction in AD is flawed, since amyloid-lowering approaches have failed to provide cognitive benefits in human clinical trials (*9*). A growing body of evidence suggests that impaired brain energy metabolism in AD may contribute to cognitive decline. At the same time, therapeutic options, such as drugs typically prescribed for metabolic disorders that improve metabolic status, may slow cognitive decline or prevent dementia progression (*10*). This is suggested by positron emission tomography imaging studies revealing baseline cerebral glucose metabolism abnormalities before the onset of cognitive symptoms in patients with AD (*11*). In addition, recent preclinical data indicate that ageing and AD are associated with the reorganization of brain energy metabolism, including an overall increase in lactate secretion and the downregulation of bioenergetic enzymes (*12, 13*).

Although current research is paving the way for developing neuroprotective therapeutics, the results of early clinical trials of drugs targeting single pathways have been mostly unsuccessful. A divergent approach combining multiple compounds that simultaneously reduce oxidative injury and improve bioenergetics, in other words, targeting various pathways has been proposed as a therapeutic strategy associated more likely with successful translational outcomes (*14*). Previous research identified limited serine availability, reduced de novo glutathione synthesis, and altered NAD+ metabolism based on the combining multi-omics profiling of transgenic AD mouse model of AD (*15*). Consistent with this, it also has been reported that age- and AD-associated metabolic shifts responded well to NAD(P)+/NAD(P)H redox-dependent reactions (*16, 17*). These findings were confirmed by human metabolomic data showing significantly altered cerebrospinal fluid (CSF) acylcarnitine levels in patients with AD, which correlated with the decline of cognitive functions and structural brain abnormalities (*18, 19*).

Based on integrative network analysis of non-alcoholic fatty liver disease multi-omics data, we have developed a mixture combined metabolic activators (CMAs) consisting of L-serine, N-acetyl cysteine (NAC), nicotinamide riboside (NR), and L-carnitine tartrate (LCAT, the salt form of L-carnitine) and showed that administration of CMAs activates mitochondria, improves inflammation markers in animals and humans (*20–24*). We have also found that the administration of CMAs promotes mitochondrial fatty acid uptake from the cytosol, facilitates the fatty acid oxidation in the mitochondria, and alleviates oxidative stress (*25*). Recently, we reported that CMAs administration effectively increased fatty acid oxidation and *de novo* glutathione generation, as evidenced by metabolomic and proteomic profiling (*20*). Moreover, plasma levels of metabolites associated with antioxidant metabolism and inflammatory proteins were improved in COVID-19 patients treated with CMAs compared to the placebo (*24*).

Here, we first analysed brain transcriptomics data obtained from 629 AD patients and 704 control subjects using genome-scale metabolic models and revealed that mitochondrial dysfunction is involved in the underlying molecular mechanisms associated with AD. Second, we tested the effect of CMAs, which has been shown to activate mitochondria in the AD rat models and showed that supplementation of the CMAs improved the AD and associated functions in animals. Next, we hypothesized that AD patients could be treated with the administration of the CMAs by activating the mitochondria in the brain tissue of the patients. Finally, we designed a randomized, double-blinded, placebo-controlled human phase 2 clinical study, studied the effect of administration on the global metabolism of AD patients and showed that administration of CMAs improves the cognitive functions in AD patients.

## RESULTS

### Analysis of transcriptomics data reveals mitochondrial dysfunction in the brain of AD patients

To identify the metabolic dysregulations based on brain transcriptomics data, we obtained the global mRNA expression profiling of 629 AD patients and 704 controls from the Religious Orders Study/Memory and Aging Project (ROSMAP) (*26–28*). We performed differential expression analysis and identified 914 significantly (p-adjusted < 1.0 E-10) upregulated and 1725 significantly (p-adjusted < 1.0 E-10) downregulated differentially expressed genes (DEGs) (Figure 1A, Dataset S1). We identified several upregulated metabolism-related genes, including pyruvate dehydrogenase kinase 4 (PDK4), carnitine palmitoyl transferase (CPT1A), hexokinase 2 (HK2), and spermine oxidase (SMOX), as well as downregulated genes including acyl-CoA dehydrogenases (ACADs), ATP synthase (ATP5MGL), and acetyl- and acyltransferases (GCNT7, MBOAT4, GALNT17). These results suggest that there may be some alterations associated with glycolysis, fatty acid biogenesis and the urea cycle in AD compared to control.

**Figure 1.**
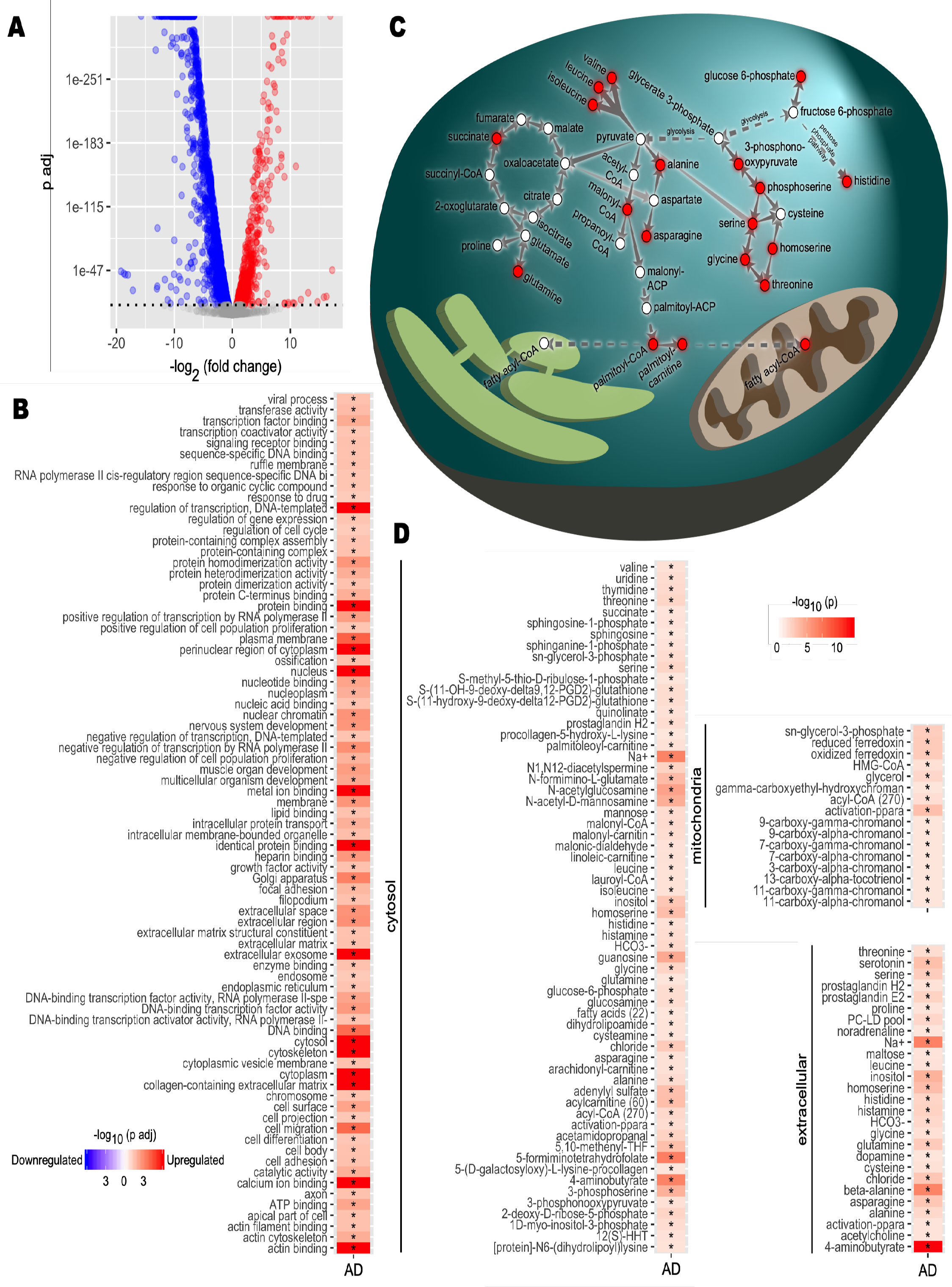
Transcriptomic analysis of brain tissue samples in ROSMAP datasets. **A)** Differential expression analysis for Alzheimer’s disease (AD) brain samples compared to healthy controls was performed. Differentially expressed genes (DEGs) were determined from gene expression values. DEGs with a p-value of 1×10-10 or smaller after Benjamini-Hochberg adjustment were determined statistically significant. Each point represents one gene. Red, significantly upregulated genes; blue, significantly downregulated genes; grey, not significant. B) Functional enrichment analysis was performed. Gene set enrichment analysis was applied on the DEGs to determine upregulated and downregulated GO terms compared to controls. Colour scale indicates direction of enrichment and p-value after Benjamini-Hochberg adjustment. Significance code: *, Adj.p< 0.05. C) Carbon metabolism pathway analysis. Alterations to pathways were inferred from reporter metabolite analysis. Key reactions and pathways linking reporter metabolites (red nodes) are shown. Reactions are simplified and arrows may represent multiple reactions. D) Reporter metabolite analysis. Statistics from DEG analysis were used to infer altered metabolites based on the iBrain2845, functional genome-scale metabolic model of brain. Asterisks indicate statistical significance based on Student’s t-test. P value <0.05.

The gene set enrichment (GSE) analysis revealed that these DEGs were significantly enriched in the protein synthesis, ATP synthesis, lipid metabolism, cell cycle, cell migration, cell differentiation and cell adhesion pathways (Figure 1B, Dataset S1). Then, we performed reporter metabolite analysis to predict the significantly changed metabolites in each subcellular compartment using a genome-scale metabolic model of brain tissue (*29, 30*). We identified numerous reporter metabolites related to glycolysis, amino acid metabolism (e.g. glycine, serine, threonine, alanine and branched-chain amino acid valine, leucine and isoleucine), mitochondrial metabolism (acyl-CoA and ferredoxin) and TCA cycle (e.g. succinate and glutamine) (Figure 1C and 1D, Dataset S1). These results suggested a widespread perturbation in energy metabolism related to brain cell survival and mitochondrial dysfunction in the brain.

### Administration of CMAs to animal models of AD

To test the effect of CMAs in animals, we provided individual metabolic activators and CMAs to the rat model of AD which has been developed after intracerebroventricular-streptozotocin (STZ) injection. We observed that administration of all constituents of CMAs (in combination or separately) significantly (p<0.05) decreased the plasma levels of triglycerides (TG) compared to the control group (Group 4; Figure 2A, Dataset S2). In parallel, administration of only serine or NR significantly decreased total cholesterol (p=0.01) and low-density lipoprotein (LDL; p=0.03) in rats (Figure 2B, Dataset S2). Additionally, a significant reduction in total cholesterol (p=0.04) and LDL (p=0.03) was observed with NAC-treated and LCAT-treated rats, respectively (Figure 2B, Dataset S2). Of note, we found significant reductions in plasma AST (p=0.03) and an increase in plasma ALP (p=0.04) concentrations only in LCAT-treated rats (Group 8) (Figure 2B, Dataset S2).

**Figure 2.**
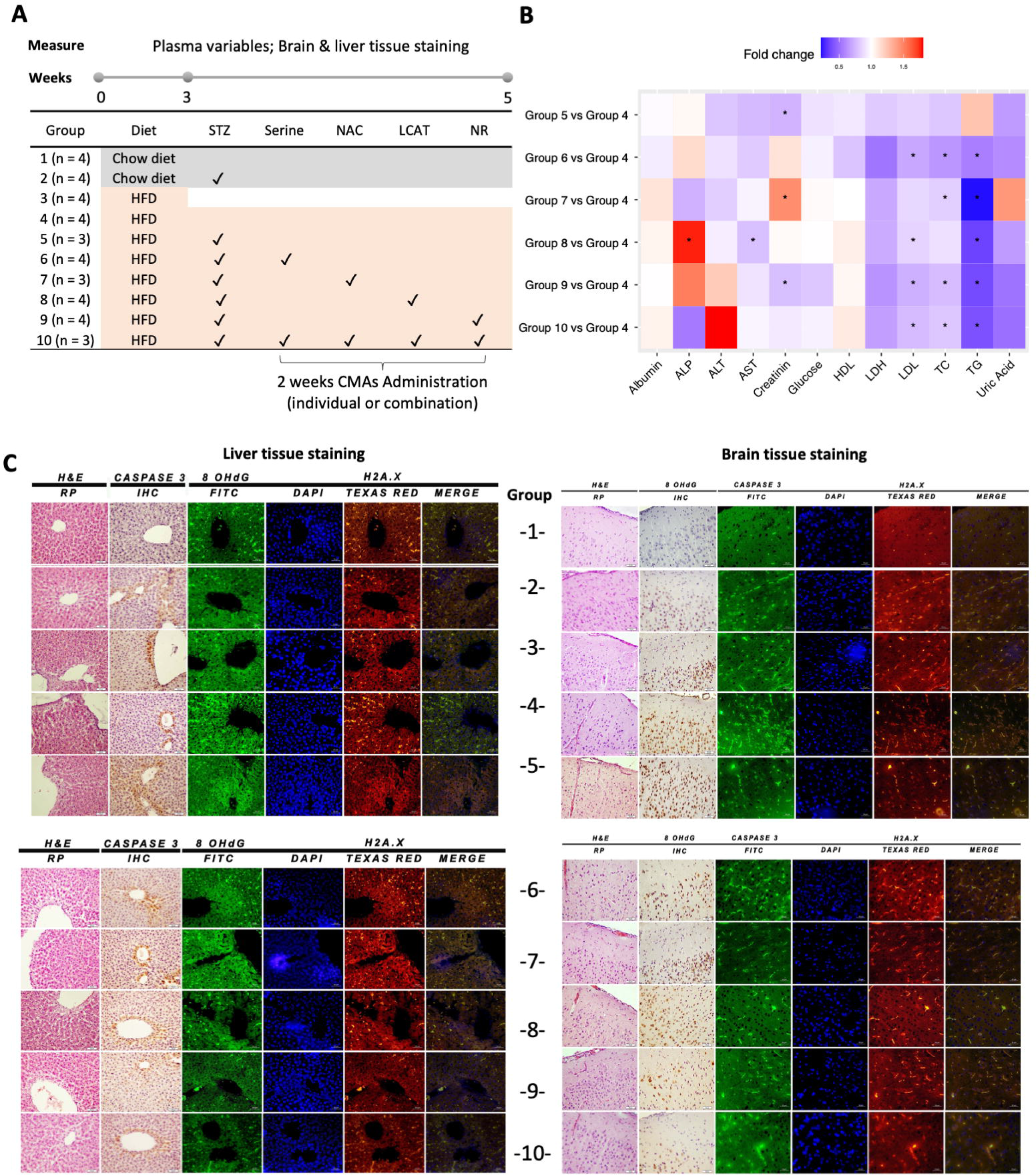
Animal study. A) Rat animal groups in in vivo experiments. Group 1 (n=4) were fed with only regular Chow diet (CHOW) for 5 weeks; Group 2 (n=4) were fed with CHOW and treated with streptozotocin (STZ). The remaining animals in Group 3 (n=4) were fed with high fat diet (HFD) for 3 weeks and sacrificed to verify model development. Groups 4-10 were fed with HFD for 5 weeks. After 3 weeks, the animals in Groups 5-10 were treated with STZ and administered with individual or combined metabolic activators, including L-serine, NAC, LCAT and NR for 2 weeks. B) Heatmap shows FC based alterations of the clinical variables in the rat study groups. Asterisks indicate statistical significance based on Student’s t-test. P value <0.05. TG, triglyceride; TC, total cholesterol; ALP, Alkaline phosphatase; AST, Aspartate aminotransferase; ALT, Alanine aminotransferase; HDL, high-density lipoprotein; LDL, low-density lipoprotein; LDH, Lactate dehydrogenase. C) Histopathological, immunohistochemical (Caspase 3) and immunofluorescence (8-OHdG and H2A.X) images of rat brain (right side) and liver tissue (left side). Slides evaluated by independent pathologist and immunopositivity scores were: Absent (-), very mild (+), mild (++), moderate (+++) and severe (++++).

Additionally, the histological analyses and immunofluorescence imaging techniques showed a significant neuronal tissue damage in the high fat diet (HFD) and HFD+STZ groups’ brains compared to those of the chow diet group (Figure 2C, Dataset S2). Specifically, the HFD groups developed more hyperaemia as well as more degeneration and necrosis in neurons (Figure 2C, Dataset S2). In parallel, DNA damage markers (namely 8-OHdG and H2A.X) and caspase 3 were elevated in the HFD groups (Figure 2C, Dataset S2). These animal models allowed us to examine each rat group’s histopathological differences and assess the brain tissue response to CMAs administration compared to the HFD+STZ group (Figure 2C, Dataset S2). We observed that hyperemia, degeneration and necrosis in neurons were improved by serine, LCAT, or NR supplementation individually as well as in combination. However, we observed a better improvement after the administration of CMAs compared to individual metabolic activators. These findings were also supported by immunohistochemical evidence of decreased immunoreactivity seen in neurons (Figure 2C, Dataset S2). Of note, rats receiving combination therapy (consisting of serine, NAC, LCAT and NR) developed less hyperaemia in brain tissue and no necrosis in neurons (Figure 2C, Dataset S2). In parallel to the improvement in the brain, we also observed dramatic improvement in the liver after the supplementation of CMAs. Scoring of histopathological, immunohistochemical and immunofluorescence findings for brain and liver tissues are presented in Table 1.

**Table 1.** Scoring histopathological, immunohistochemical and immunofluorescence findings in brain tissues and liver tissues

| Organ | Groups | Hyperemia | Degeneration in Neurons | Necrosis in Neurons | 8-OHdG (IHC) | Caspase 3 (IFA FITC) | H2A.X (IFA Texas Red) |
| --- | --- | --- | --- | --- | --- | --- | --- |
| BRAIN | CHOW | - | - | - | - | - | - |
|  | CHOW + STZ | +++ | +++ | +++ | +++ | +++ | +++ |
|  | HFD 3w | +++ | +++ | +++ | +++ | +++ | +++ |
|  | HFD 5w | ++++ | ++++ | ++++ | ++++ | ++++ | ++++ |
|  | HFD+STZ | ++++ | ++++ | ++++ | ++++ | ++++ | ++++ |
|  | HFD+STZ+Serine | +++ | +++ | ++ | +++ | +++ | +++ |
|  | HFD+STZ+NAC | ++++ | ++++ | +++ | +++ | +++ | +++ |
|  | HFD+STZ+LCAT | +++ | +++ | ++ | +++ | +++ | +++ |
|  | HFD+STZ+NR | +++ | +++ | ++ | +++ | +++ | +++ |
|  | HFD+STZ+Serine+NAC+LCAT+NR | ++ | ++ | + | ++ | ++ | ++ |
| LIVER | CHOW | - | - | - | - | - | - |
|  | CHOW + STZ | +++ | +++ | +++ | +++ | +++ | +++ |
|  | HFD 3w | +++ | +++ | +++ | +++ | +++ | +++ |
|  | HFD 5w | ++++ | ++++ | ++++ | ++++ | ++++ | ++++ |
|  | HFD+STZ | ++++ | ++++ | ++++ | ++++ | ++++ | ++++ |
|  | HFD+STZ+Serine | +++ | +++ | +++ | +++ | +++ | +++ |
|  | HFD+STZ+NAC | +++ | ++++ | +++ | +++ | +++ | +++ |
|  | HFD+STZ+LCAT | +++ | +++ | +++ | +++ | +++ | +++ |
|  | HFD+STZ+NR | +++ | +++ | ++ | +++ | +++ | +++ |
|  | HFD+STZ+Serine+NAC+LCAT+NR | +++ | +++ | + | + | ++ | ++ |

### CMAs Improves Cognition and Blood Parameters in Alzheimer’s Disease Patients

To test the effect of the CMAs in AD patients, we performed a double-blind, randomized, placebo-controlled phase 2 study and screened 89 adults diagnosed with AD. We recruited 69 patients older than 50 years with mild to moderate AD according to ADAS-Cog (AD Assessment Scale-cognitive subscale; ADAS≥12) and the Clinical Dementia Rating Scale Sum of Boxes (CDR-SOB; CDR≤2). Of the 69 patients, 47 were randomly assigned to the CMA group and 22 to the placebo group and completed visit 2 after 28 days. Of these patients, 60 (40 in the CMA group and 20 in the placebo group) completed visit 3 after 84 days (Figure 3A, Dataset S3). We assessed the clinical variables on Days 0, 28 and 84, and analysed the differences between the time points in the CMA and placebo groups (Dataset S4).

**Figure 3.**
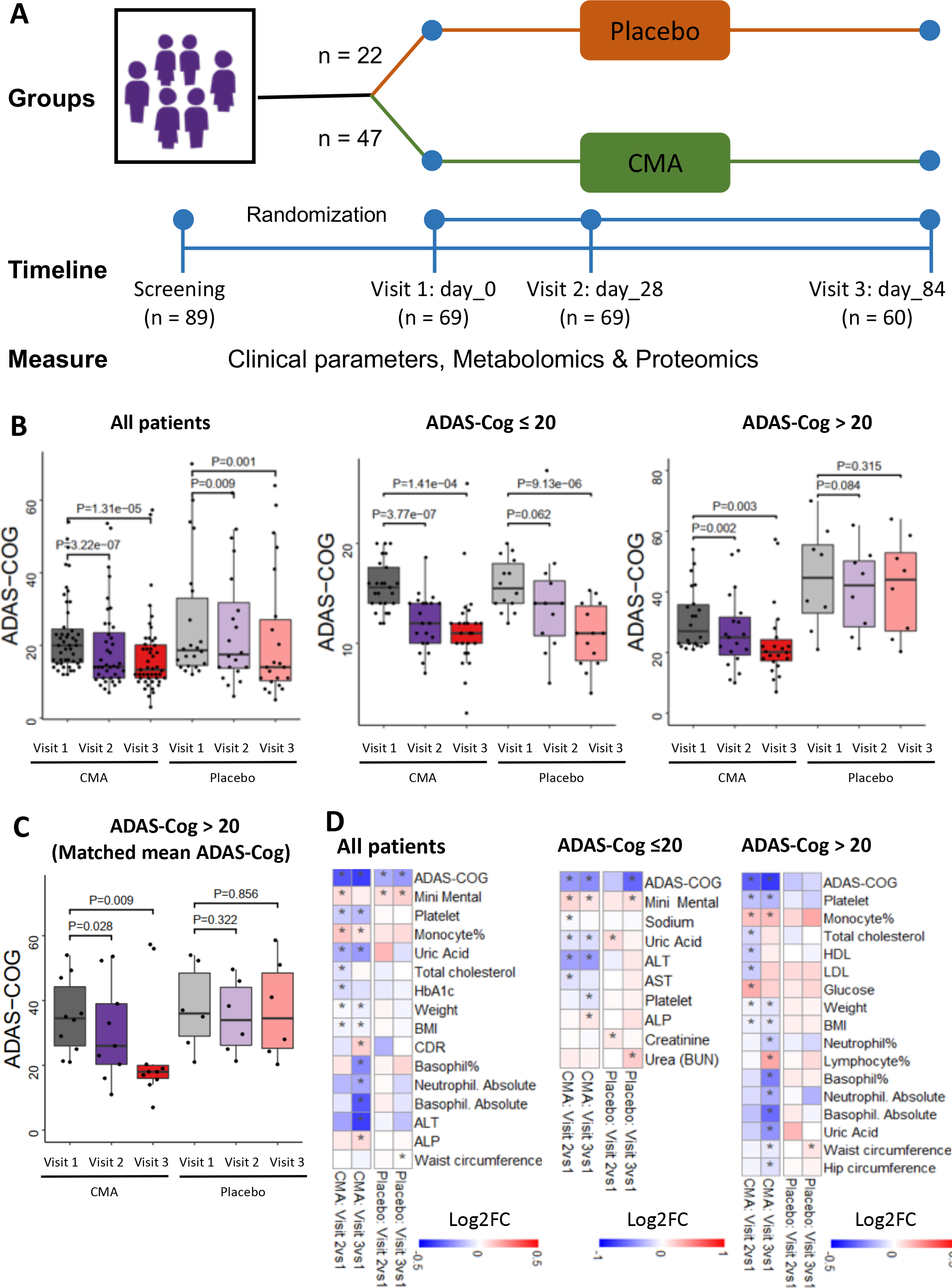
CMAs Improves ADAS-Cog scores and clinical parameters. A) Study design for testing the effects of CMAs in AD patients. B) Differences in ADAS-Cog scores in the CMA and placebo groups on Days 0, 28 and 84 are presented. Additionally, differences in ADAS-Cog scores were analysed by stratifying the patients into high and low levels of ADAS-Cog groups (> 20 ADAS-Cog is high, ≤ 20 is low). As decreased ADAS-Cog score is the indicator of the improved cognitive function in AD patients, the ADAS-Cog scores is significantly decreased on Day 28 vs Day 0 (Log2FoldChange (FC)= -0.33, (26% improvement), p-value=0.0000003) and Day 84 vs Day 0 (Log2FC= -0.37, (29% improvement), p-value=0.00001) in the CMA group. A slightly but significant decrease is found in the placebo group on Day 28 vs Day 0 (Log2FC= -0.16, (12% improvement), p-value=0.009) and Day 84 vs Day 0 (Log2FC= -0.19, (14% improvement), p-value=0.001) due to the recommendations of exercise and Mediterranean diet to all AD patients participated in the trial. The differences between clinical parameters have also been analysed by stratifying the patients into low-scored (mild patients) and high-scored (severe patients) ADAS-Cog groups (>20ADAS-Cog score is high, ≤ 0 is low). The ADAS-Cog scores is significantly decreased on Day 28 vs Day 0 (Log2FC= -0.31, (24% improvement), p-value=0.002) and Day 84 vs Day 0 (Log2FC=-0.38, (30% improvement), p-value=0.003) in the high scored CMA group and no significance difference in the high-scored placebo (p>0.05 in both time points) group. C) We selected 10 patients from the severe (ADAS-COG > 20) CMA group with matched ADAS-COG values to the placebo group (P-Value: 0.693) and presented the ADAS-Cog scores. We recalculated the differences in ADAS-COG scores, and found significant improvement in the CMA group whereas no significant difference in the placebo group. D) Heatmaps shows log2FC based alterations of the clinical variables, which are compared before and after the administration of CMA in both drug and placebo groups. Asterisks indicate statistical significance based on Student’s t-test. P value <0.05. Log2FC: log2(fold change).

The patients’ mean age participated in the study was 70.8 years (56-86 years), and 52.1 % were men (Dataset S4). The baseline demographic and clinical characteristics were similar in the CMA and placebo groups (Dataset S4). Regarding safety, no severe adverse events occurred and 4 patients (5.8 %) reported adverse events. All had a mild rash on the body and decided to complete the study (Dataset S3).

We measured clinical variables in all patients and analysed the differences before and after administration in the active and placebo groups (Figure 3B, Dataset S4). As decreased ADAS-Cog score is the indicator of the improved cognitive function in AD patients, the ADAS-Cog scores is significantly decreased on Day 28 vs Day 0 (Log2FoldChange (FC)= -0.33, (26% improvement), p-value=0.0000003) and further decreased on Day 84 vs Day 0 (Log2FC= - 0.37, (29% improvement), p-value=0.00001) in the CMA group. A slightly but significant improvement is also found in the placebo group on Day 28 vs Day 0 (Log2FC= -0.16, (12% improvement), p-value=0.009) and Day 84 vs Day 0 (Log2FC= -0.19, (14% improvement), p-value=0.001) due to the recommendations of exercise and Mediterranean diet to all AD patients participated in the trial.

We also analysed the differences between clinical parameters by stratifying the patients into low-scored (mild patients) and high-scored (severe patients) ADAS-Cog groups (>20ADAS-Cog score is high, ≤20 is low). More interestingly, we found a significant improvement of ADAS-Cog scores between Day 28 vs Day 0 (Log2FC= -0.31, (24% improvement), p-value=0.002) and Day 84 vs Day 0 (Log2FC=-0.38, (30% improvement), p-value=0.003) in the severe CMA group and no significance difference in the severe placebo (p>0.05 in both time points) group (Figure 3B, Dataset S4). As shown on Figure 3B, we observed a significant difference in the baseline value distribution and mean of ADAS-Cog scores in the severe (ADAS-COG > 20) CMA and placebo group due to the randomization of the subjects. To verify our results, we selected 10 patients from the CMA group with matched ADAS-COG values to the placebo group (P-Value: 0.693) and presented the ADAS-Cog scores in Figure 3C. We recalculated the differences in ADAS-COG scores, and again found significant improvement in the CMA group, whereas no significant difference in the placebo group. Our results indicated that the AD patients with high ADAS-Cog scores are more responsive to CMA.

Analysis of secondary outcome variables showed that serum alanine aminotransferase (ALT) levels (Log2FC=-0.38, (30% improvement), p-value=0.01) and the uric acid levels (Log2FC=-0.19, (14% improvement), p-value=0.001) were significantly lower on Day 84 vs Day 0 only in the CMA group (Figure 3D, Dataset S4). This reduction was seen both in high- and low-ALT level groups. In contrast, we found that no significantly altered parameters on Day 84 vs Day 0 in the placebo group (Figure 3D, Dataset S4). Hence, we observed that the ALT level and uric acid were significantly improved due to the administration of CMAs as previously reported in the Phase 2 NAFLD and Phase 3 Covid-19 clinical trials (*24, 31*).

We also measured the level of complete blood count parameters and found that their levels were significantly changed in the CMA group (Figure 3D, Dataset S4). We found that the levels of platelets, basophil% and absolute numbers of basophil and neutrophil were significantly lower on Day 84 vs Day 0 only in the CMA group. In contrast, we found that the levels of monocytes were significantly increased on Day 84 vs Day 0 in the CMA group (Figure 3D, Dataset S4). Hence, our analysis indicated that the administration of CMAs improved the clinical parameters in parallel to the improvement in cognitive functions in AD patients.

### Blood profile informs the response to CMAs

Treatment response variability and clinical heterogeneity in AD are well documented in the literature. We observed interindividual variability in responses to CMAs as well as in clinical measures. Therefore, we hypothesized that some of the patients would respond better to CMAs than others and that clinical measurements could define these subsets.

To determine whether alanine transferase (ALT), a marker for liver damage, could indicate a better response to CMAs, we stratified the patients into high and low ALT groups by the median value of ALT of all patients on Day 0. As shown in Figure 4A, the patients of the CMA group with low ALT achieved a significant improvement in ADAS-Cog score over different time points, while the patients in the placebo group had no improvement. In contrast, the patients of the CMA group with high ALT levels also exhibited an improved (i.e., decreased) ADAS-Cog score, but the degree of change was not as much as the patients in the CMA group with low ALT levels. Moreover, patients in the placebo group with high ALT levels also achieved an improved ADAS-Cog score. Thus, these results suggest that the patients with low ALT levels are more responsive to CMAs.

**Figure 4.**
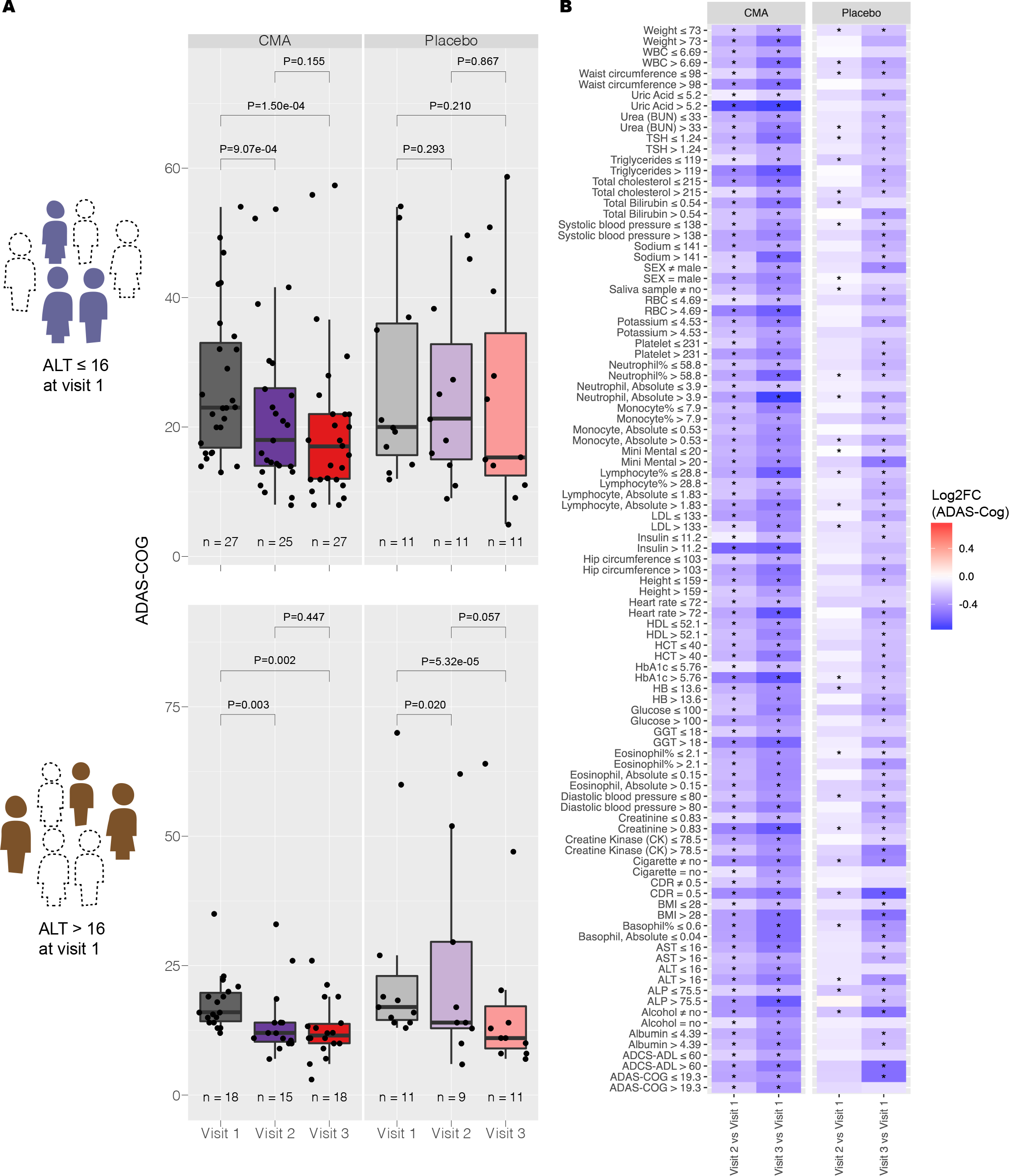
Identification of clinical variables informative for response to CMAs administration. A) Distribution of ADAS-Cog scores over visit number for patients with ALT ≤ 16 IU/L at visit 1 (upper panel) and patients with ALT > 16 IU/L at visit 1 (lower panel). B) Between-visit changes to ADAS-Cog with various clinical variable groupings are shown. Only those clinical variable groupings resulting in a more significant change to ADAS-Cog in CMA group compared to placebo group (a p-value of 0.05 or better are shown). Colour scale indicates log2 fold change to ADAS-Cog between visits. Statistical significance between visits was determined by a paired t-test across individuals who attended both visits. Asterisks indicate statistical significance p< 0.05.

We repeated this stratification for each blood parameter to determine the patient conditions in which CMAs produces the greatest response (Figure 4B). In addition to low ALT, we identified high ALP, low GGT, high HCT, high HbA1c, high insulin, high uric acid, high ADAS-Cog, high basophil count, and high red blood cell count as indicators for better responsiveness to CMAs. We also found that individuals who do not drink alcohol or smoke also respond better to CMAs.

### CMAs Increases the Plasma Levels of Metabolites Associated with Metabolic Activators

We first analysed the plasma levels of serine, carnitine, NR and cysteine, and their by- products. CMAs administration increased the plasma levels of metabolic activators on Day 84 vs Day 0 in the CMA group (Figure 5A, Dataset S5). Moreover, the plasma levels of NR, 1-methylnicotinamide, nicotinurate, N1-methyl-2-pyridone-5-carboxamide and nicotinamide (associated with NR and NAD+ metabolism); of serine, glycine and sarcosine (associated with serine and glycine metabolism); and of deoxycarnitine and carnitine (associated with carnitine metabolism) were significantly higher in the CMA group on Day 84 compared to Day 0.

**Figure 5.**
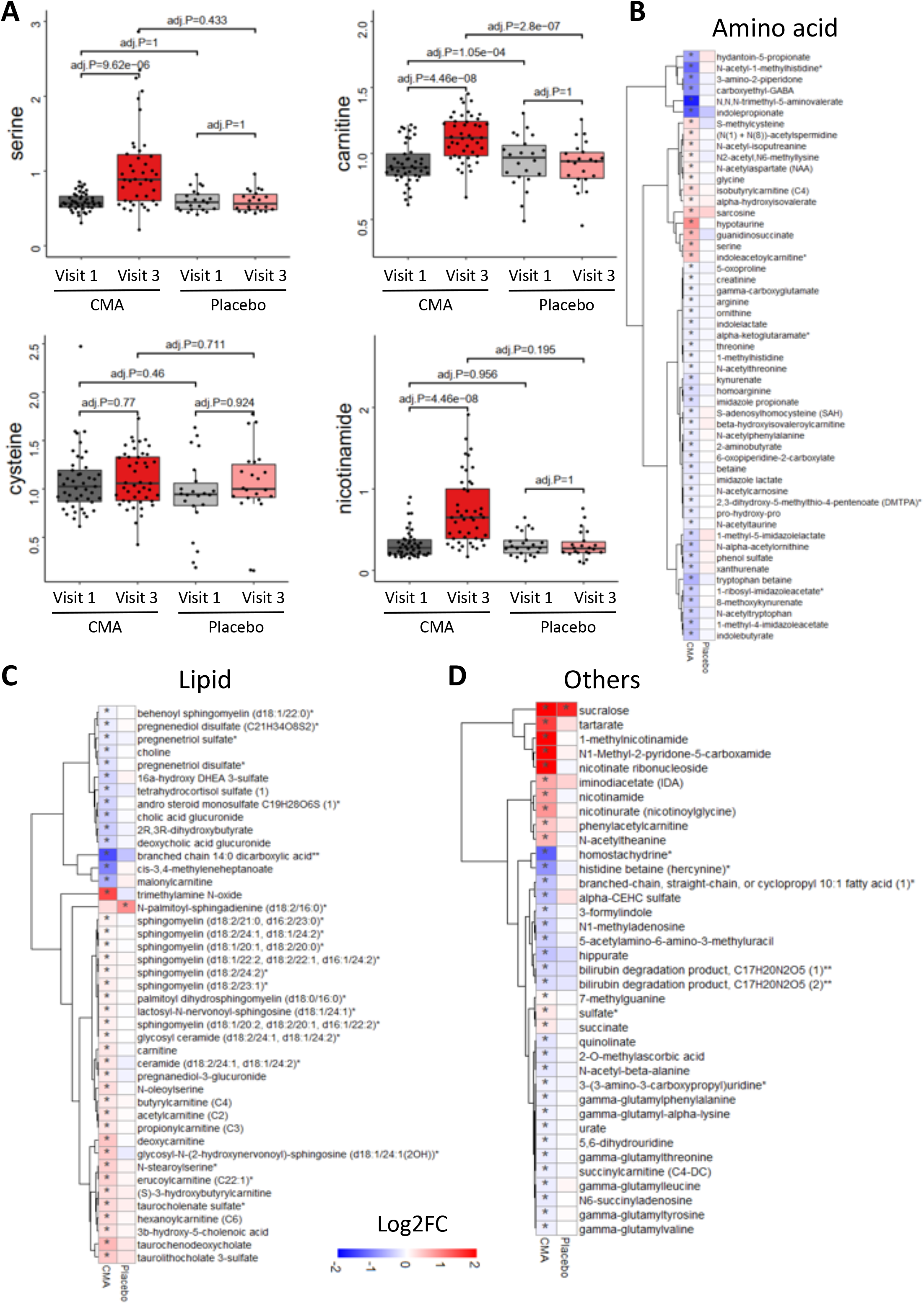
CMAs alters plasma metabolite levels. A) Differences in the plasma levels of individual CMAs including serine, carnitine, cysteine and nicotinamide are shown in the CMA and placebo groups on Days 0 and 84. Plasma level of B) amino acids, C) lipids and D) other metabolites that are significantly different between Day 84 vs Day 0 in the CMA and placebo groups are presented. Adj. p< 0.05. Heatmap shows log2FC values of metabolites between Day 84 vs Day 0. Asterisks indicate statistical significance based on paired Student’s t test. Adj.p< 0.05. Log2FC: log2(fold change).

Next, we investigated the relationship with the plasma level of administrated metabolic activators and other metabolites. We analysed 195 of the most significantly correlated plasma metabolites with serine, L-carnitine, NR, and cysteine (Dataset S6). We found two clusters of metabolites that are significantly correlated with cysteine only or together with serine, carnitine and NR. We observed that cysteine had a different plasma changes compared to the other three metabolic activators as has been reported in previous clinical trials (*24, 31*).

### Effect of CMAs on Global Metabolism

We identified the significantly (FDR<0.05) different plasma metabolites on Day 84 vs Day 0 and found that the plasma levels of 132 metabolites were significantly different in the CMA group (Figure 5, Dataset S5). Evaluation of plasma metabolites that differed significantly on Day 84 vs Day 0 in each group showed that a larger number of metabolites related to amino acid (n=53), lipid metabolism (n=42) and other metabolic pathways (n=37) were altered in the CMA group compared to the placebo group (Figure 5, Dataset S5).

N-acetyl aspartate (NAA) is one of the most abundant brain metabolites and its reduced plasma levels are associated with brain tissue damage. Previous research revealed the importance of NAA to maintain energy metabolism in the central nervous system (*32*). In our study, we observed that plasma levels of NAA significantly increased on Day 84 vs Day 0 in the CMA group (Figure 5B, Dataset S5). Another upregulated metabolite on Day 84 vs Day 0 in the CMA group is sarcosine (a derivative of glycine) which has been widely studied for its improving effects of cognitive symptoms by different pharmacological activities in neurons (*33*). Of note, quinolinic acid (an endogenous excitotoxin acting on N-Methyl-D-aspartate receptors leading to neurotoxic damage) levels significantly decreased on Day 84 vs Day 0 only in the CMA group (Figure 5B, Dataset S5).

Increased plasma homocysteine levels is a known risk factor for AD, and several animal studies implicated the promising results of methionine restriction (*34, 35*). In our clinical trial, plasma levels of S-adenosylhomocysteine as well as 2,3-dihydroxy-5-methylthio-4-pentenoate (DMTPA) and N-acetyl taurine were significantly downregulated on Day 84 vs Day 0 in the CMA group (Figure 5B, Dataset S5). Of note, reductions of these metabolites are significantly correlated with serine and NR supplementation (Figure 6A, Dataset S6).

**Figure 6.**
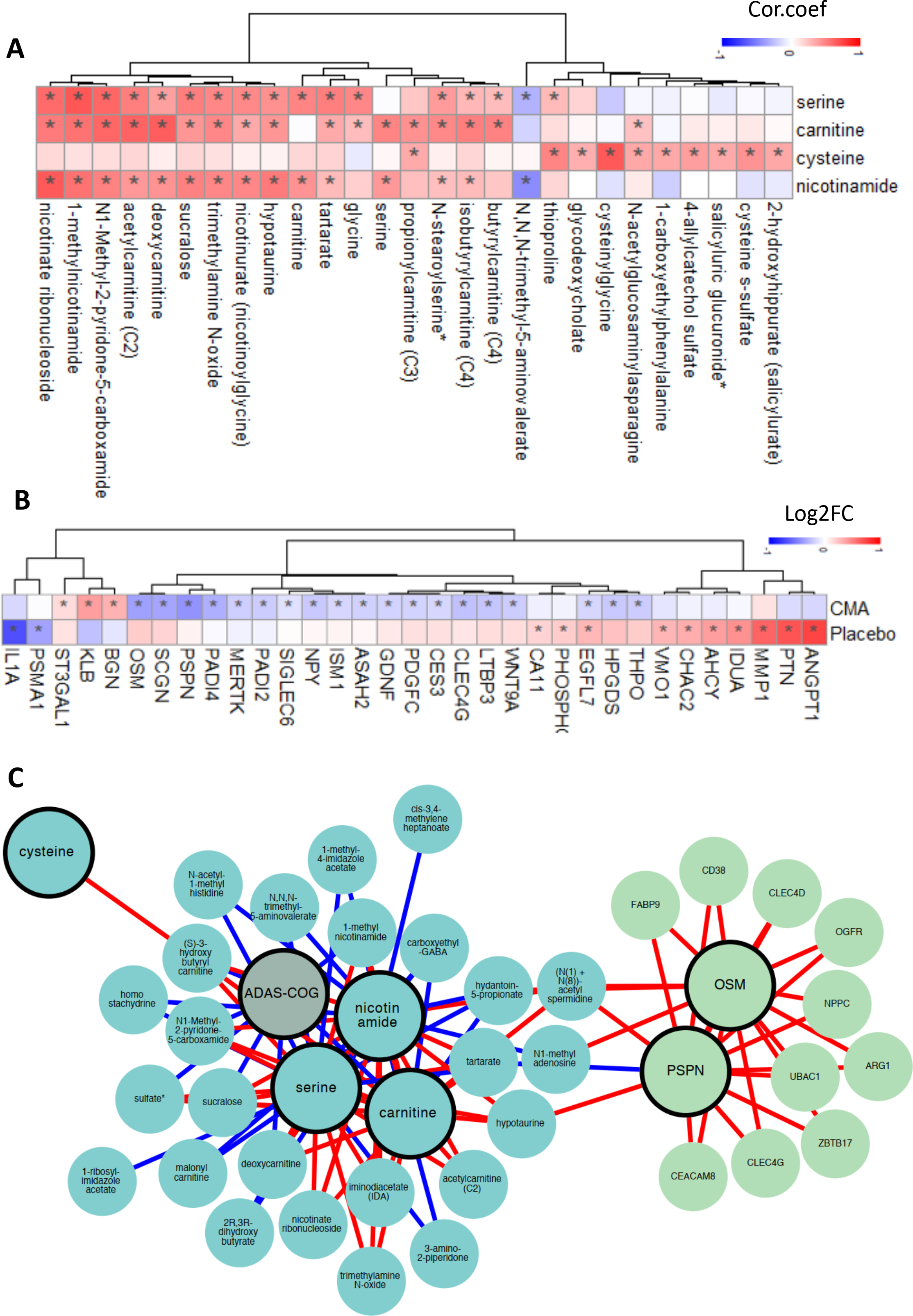
Correlation of CMAs with plasma metabolites and altered plasma protein levels. A) Associations between the plasma level of individual CMAs and the 10 most significantly correlated plasma metabolites are presented. Asterisks indicate statistical significance (Adj.p < 0.05) based on Spearman correlation analysis. Cor.Coeff: Correlation coefficient B) Heatmap shows log2FC based alterations between the significantly different proteins on Day 84 vs Day 0 in the CMA and placebo groups. Asterisks indicate statistical significance based on paired Student’s t test. p < 0.01. C) Integrated multi-omics data based on network analysis represents the neighbours of the CMAs, including serine, carnitine, nicotinamide and cysteine, and ADAS-Cog scores. Only analytes that are significantly altered in CMA Day 84 vs Day 0 are highlighted.

Increased plasma levels of metabolites in the kynurenine pathway are associated with AD severity (*34*). In our study, we found that plasma levels of kynurenate and 8-methoxykynurenate were significantly lower on Day 84 vs Day 0 in the CMA group (Figure 5B, Dataset S5). Reduction in the plasma level of kynurenate was positively correlated with plasma serine levels (Dataset S6). Kynurenate, which has a prooxidant effect, is the product of the tryptophan degradation pathway. Its aerobic irradiation produces superoxide radicals and leads to cytochrome C reduction (*36*). It has been reported that increased levels of kynurenine lead to cell death through the reactive oxygen species (ROS) pathway in nature killer (NK) cells (*37*) and lower blood pressure in systemic inflammation (*38*).

Emerging evidence indicates a link between the abnormal kidney function and AD, but the potential impact of kidney on cognitive impairment is still undetermined (*39*). Recent studies showed that plasma levels of N,N,N-trimethyl-5-aminovalerate are involved in lysine metabolism, and serve as an indicator of elevated urinary albumin excretion (*40*). Here, we found that the plasma level of N,N,N-trimethyl-5-aminovalerate was significantly decreased on Day 84 vs Day 0 in the CMA group (Figure 5B, Dataset S5) and significantly inversely correlated with the plasma level of serine and NR. Moreover, the plasma level of creatinine was also significantly decreased on Day 84 vs Day 0 in the CMA group (Figure 5B, Dataset S5). The plasma reduction on creatinine is inversely correlated with the plasma level of serine (Dataset S6). Additionally, our analysis revealed decreased levels of several metabolites belonging to histidine metabolism in the CMA group on Day 84 vs Day 0. Among those N-acetyl-1-methylhistidine is associated with chronic kidney disease and showed a significant negative correlation with serine supplementation (Figure 5B, Dataset S6). Also, we found that plasma levels of metabolites related to the urea cycle (3-amino-2-piperidone, arginine, homoarginine, N-alpha-acetylornithine, ornithine and pro-hydroxy-pro) were significantly decreased in the CMA group on Day 84 vs Day 0 (Figure 5B, Dataset S5) and inversely correlated with the plasma level of serine and NR (Dataset S6).

Lipids play a fundamental role in the pathophysiology of neurodegenerative diseases, including AD. Specific lipid species of cellular membranes (e.g., cholesterol and sphingolipids) are not only structural components of cell membranes but also regulate a plethora of critical aspects of brain functions(*41*). In our study, plasma levels of a considerable number of metabolites associated with sphingomyelins and fatty acid metabolism (acyl carnitines) were significantly increased on Day 84 vs Day 0 in the CMA group (Figure 5C, Dataset S5). Interestingly, plasma levels of pregnenolone steroids and 2R,3R-dihydroxybutyrate were significantly decreased on Day 84 vs Day 0 (Figure 5C, Dataset S5). These alterations were significantly positively correlated with carnitine and serine levels (Dataset S6).

### Effect of CMAs on Plasma Proteins

Plasma levels of 1466 protein markers were measured with the plasma proteome profiling platform Proximity Extension Assay quantifying the plasma level of target proteins. After quality control and exclusion of proteins with missing values in more than 50% of samples, 1463 proteins were analysed (Dataset S7). Proteins whose levels differed significantly between the visits in the CMA and placebo groups are listed in Dataset S7.

We analysed the effect of CMAs on plasma protein profile and found that 22 proteins were significantly (p-value<0.01) different in the CMA group on Day 84 vs Day 0. Nineteen of these proteins were significantly decreased, whereas 3 of these proteins were significantly increased on Day 84 vs Day 0. After filtering out the proteins based on log2FC, we found that the plasma levels of PSPN, OSM, PADI4, PDGFC, SCGN, LTBP3, CLEC4G, MERTK, WNT9A, ISM1, ASAH2, CES3, HPGDS, NPY, THPO, SIGLEC6, GDNF, PADI2 and EGFL7 were significantly downregulated in the CMA group. The plasma level of KLB, BGN, and ST3GAL1 was significantly upregulated in the CMA group (Figure 6B, Dataset S7). We observed that only 1 significantly (p-value<0.01) altered protein - EGFL7 upregulated- in the placebo group (Figure 6B, Dataset S7). Hence, we observed that plasma levels of proteins are significantly altered in response to CMA treatment.

The proteomic analysis in this study revealed significant alteration in levels of several critical proteins that play an essential role in the pathogenesis of AD. For instance, levels of MertK (*42, 43*), EGFR (*44, 45*), oncostatin (*46–50*), PAD4 (*51, 52*), LTGF (*53–57*), and TPO (*58*), known as a strong inducer of neuro-inflammation, amyloid production and apoptosis, decreased. In contrast, proteins with neuroprotective and pro-cognitive properties, such as Klotho (*59, 60*) and ST3GAL1 (*61*), increased after CMA treatment. More interestingly, the majority of the analysed proteins were also found to be significantly altered in recent human AD studies (*53-57, 62-68*). KlothoB levels were also significantly altered after CMA treatment, consistent with their neuroprotective role as a cofactor and neurotrophic factor. In this context, recent studies have shown that KlothoB indirectly regulates glucose and energy metabolism through F2F1, expressed in certain areas of the brain involved in learning and memory (*67*). Moreover, GABA signalling has also been shown to play a critical role in mediating the detrimental effects of increased dihydroxybutyrate levels in the progression of MCI (*69*). Interestingly, our metabolomic study indicated decreased post-therapeutic dihydroxybutyrate levels. Although the exact pathways involved in the metabolic generation of DHBA are still far from clear, it has been hypothesized that dihydroxybutyrate levels may be a compensatory response to increased cellular stress secondary to compromise of the Krebs cycle function, creating an alternative energy production pathway in AD (*69*). This represents indirect evidence to suggest that our treatment exhibits an energetic regulatory function.

### Integrative Multi-Omics Analysis

Multi-omics data integrations have been proven to give novel insights and a more holistic view of the human body, in both healthy and disease states (*70*). In this study, we generated an integrative multi-omics network using metabolomics and proteomics data, coupled with detailed clinical variables, to understand the functional relationships between analytes from the same and different omics data types. We generated the network using the method used in iNetModels (*71*), to which we also deposited our network. The network consists of 937,282 edges from 2,273 nodes (36.3% network density, Dataset S8).

We extracted a sub-network to highlight the interactions between the individual metabolic activators, cognitive function (ADAS-Cog scores), two highlighted proteins (OSM and PSPN), and their top neighbours (Figure 6C). From the sub-network, ADAS-Cog was shown to be negatively associated with carnitine (and its derivatives) and nicotinamide associated metabolites, where the metabolic cofactors were negatively associated with fatty acid and histidine metabolism. Finally, we observed that, among others, OSM and PSPN were positively associated with immune and cell cycle-related proteins.

Subsequently, we performed centrality analysis to identify the most central analytes in the networks. The top 20 most central metabolites were dominated by amino acid metabolites (tryptophan, glutamate, and branched-chain amino acid metabolism) and lipid metabolites (androgenic steroid pathway), where top proteins were related to, among others, short- and long-term memory (CALB1), lipid metabolism (PLA2G10), and immune response (SELPLG, CLEC4D, and LGALS7).

Furthermore, we performed community analysis within the network using the Leiden algorithm. We discovered 3 modules that showed significant interaction among the members. In cluster-0, the biggest clusters, the top nodes were related to tryptophan metabolism (indoleacetate), fatty acid metabolism (3-hydroxyoctanoate), and steroid metabolism (11-ketoetiocholanolone glucuronide and 11-beta-hydroxyetiocholanolone glucuronide). Moreover, we found 2 top proteins in the same cluster, ACTA2 and IGFBP1, that have been associated with AD (*72, 73*). In cluster-1, the top nodes were associated with leucine metabolism (3-hydroxy-2-ethylpropionate), ceramide phosphatidylethanolamine, and a carnitine metabolite (erucoylcarnitine), meanwhile, cluster-2’s central nodes were related to methionine metabolism and aminosugar metabolism (N-acetylglucosamine/N-acetylgalactosamine). These results showed that the integrative multi-omics network analysis can be used to strengthen the results from single omics analyses and identify key analytes associated with AD. Moreover, it provided new insights by elucidating the functional relationships within and between different omics data.

In evaluating the correlations between each cofactor (used in the present study for therapeutic purposes) and clinical, metabolic, and proteomic parameters, we identified significant correlations between serine, carnitine, cysteine, and nicotinamide levels and improved peripheral blood parameters, such as liver function, CBC, and glycated hemoglobin (HbA1c), which are relevant to the pathogenesis of AD. Accordingly, improved ADAS-Cog scores were also associated with changes in serum serine and carnitine, which fit well with their well-known pro-cognitive and energy-boosting effects. Similar results were also observed for metabolomic and proteomic data. The majority of the cofactors exhibited significant correlations with improved metabolites and proteins (either increased or decreased) relative to a slower degeneration process in AD. It is worth mentioning here that that two of the proteins, OSM and PSPN, most strongly associated with other beneficial protein metabolites, were also related to several critical amino acid alterations, such as spermidine and hypotaurine, which may suggest a metabolic shift from the protein to amino acid metabolism to compensate the energy deficit reported in AD.

## DISCUSSION

Here, we show that oral administration of CMAs has profound effect on cognitive function after only 84 days of treatment in AD patients based on ADAS-Cog scores. We showed that cognitive functions in AD patients is improved 29% in the CMA group whereas 14% in the placebo group after 84 days of CMAs administration. We also recently tested the effect of CMAs administration on an independent cohort of Parkinson’s disease patients in a Phase 2 clinical trial, and found that oral CMAs administration has a profound effect on cognitive function without altering motor scores in Parkinson’s Disease (*74*). As the increased Montreal Cognitive Assessment (MoCA) scores is known as the indicator of the increased cognitive functions in PD patients, we observed that the mean MoCA scores were significantly higher in the CMA group both on Day 28 vs Day 0 (log2FoldChange(FC)=0.17, (13% improvement), p=0.001) and on Day 84 vs Day 0 (log2FC=0.27, (21% improvement), p= 0.0001). We also observed a significant increase on MoCA scores in placebo group on Day 28 vs Day 0 (log2FC=0.16, (12% improvement), p=0.001) and on Day 84 vs day 0 (log2FC=0.15, (11% improvement), p=0.04) due to the recommendations of exercise and Mediterranean diet to all PD patients participated in the trial. Notably, the degree of increase of MoCA was much higher on Day 84 vs Day 0 in the CMA group than in the placebo group, suggesting the PD patients benefitted from CMA treatment after 84 days of treatment. Even though, Montreal Cognitive Assessment (MoCA) scores have been used to evaluate the cognitive function in the PD patients, the effect of CMAs administration on cognitive function have been verified in an independent patient group with different neurological diseases.

Clinically, when AD patients were stratified by high and low ADAS-Cog scorer, we observed that patients with lower ADAS-Cog scores in the placebo group also showed improved cognitive function similar to the CMA group. However, for the patients with higher ADAS-Cog scores, cognitive function was not improved in the placebo group, while a positive effect was observed for the patients treated with metabolic activators. This is particularly interesting, since this patient group lacks current therapeutic regimes, except for palliative support. Apart from clinical severity, we also observed that various clinical variables were also related to the treatment response. For example, patients with low ALT, who did not drink alcohol or smoke, and who had signs of an increased metabolic load (i.e., increased HbA1c and insulin levels) or impaired CBC values responded better to treatment. Additionally, the treatment significantly improved the altered metabolic and CBC parameters described above.

The effect of oral administration of CMAs was substantiated with a comprehensive analysis of protein and metabolites in the plasma of the patients using a multi-omics analytical platform. The clinical results are consistent with the genome-scale metabolic modelling of more than 600 AD patients showing evidence of mitochondrial dysfunction. It is also consistent with the results from an animal model demonstrating improved AD-associated histological parameters in animals treated with an oral administration of CMAs. Thus, the present study suggests an attractive therapeutic regime for improving mitochondrial dysfunction in AD patients.

The metabolomics data confirmed the expected biological outcomes of CMA treatment. Levels of plasma nicotinamide and related metabolites increased, suggesting that NR provided sufficient substrate for mitochondrial fatty acid oxidation. In addition to its role as a cellular metabolite, NAD+ functions as an essential cofactor for the DNA repair protein PARP1 (*17*). Hyperactivation of PARP1 and decreased NAD+ have been already identified in the brains of patients with AD (*75, 76*). Serine plasma levels also increased, suggesting that CMA treatment improves the serine deficiency associated with AD. For instance, a recent study showed that the adenosine triphosphate (ATP)-reducing the effect of glucose hypometabolism was restored with oral serine supplementation, suggesting the potential use of oral serine as a ready-to-use therapy for AD (*77*). The exact mechanism of action also applies to cysteine. As a glutathione precursor, cysteine is even more specific in acting as an antioxidant and anti-inflammatory agent, maintaining the mitochondrial energetic homeostasis and key neurotransmitter systems, such as glutamate, involved in learning and memory (*78, 79*). Accordingly, NAC has been tested as a medication in AD and found to exhibit effects suggestive of future potential use as an alternative medication (*80*). More importantly, fatty acid oxidation and carnitine metabolism were significantly facilitated, as shown by the robust increase in plasma levels of carnitine. These findings fit well with recent human data showing that severe disturbances in carnitine metabolism frequently occur in individuals with AD, in association with severe mitochondrial dysfunction (*81, 82*). Cristofano et al. showed a progressive decrease in carnitine serum levels in individuals shifting from normal status to AD, suggesting that decreased serum concentrations of carnitine may predispose to AD (*83*). In support of this hypothesis, human clinical studies have demonstrated the pro-cognitive effects of carnitine in mild cognitive impairment and AD(*84–86*). This, in turn, led to the suggestion that stabilizing the bioenergetic balance may slow or even reverse mild cognitive impairment and the progression of dementia in patients with AD.

In addition, the levels of tryptophan metabolites, including kynurenate, kynurenine, and tryptophan betaine, decreased significantly after CMA treatment. Increased levels of these metabolites were previously shown to be associated with increased neurodegeneration and clinical cognitive impairment through an increased oxidative load and the formation of neurofibrillary tangles (NFTs) (*87, 88*). For instance, recent data showed a synergistic relationship between β-amyloid 1-42 and enzymatic activations of the tryptophan kynurenine pathway, resulting in increased oxidative stress, which may be associated with the formation of NFTs and senile plaque development (*89*). Also, one recent study revealed that tryptophan-2,3-dioxygenase (TDO) was highly expressed in the brains of patients with AD and co-localized with quinolinic acid, NFTs, and amyloid deposits in the hippocampus of post-mortem brains of patients with AD (*90*).

We also observed significantly increased levels of NAA, sarcosine, methionine, cysteine, and S-adenosylmethionine (SAM) and decreased levels of histidine, tryptophan quinolate, and urea cycle metabolites, which play a critical role in cognitive and mitochondrial functions. For instance, increased NAA may provide an additional energy source for intercellular metabolite trafficking during the neurodegenerative process, especially when glucose metabolism is downregulated (*32*). Similarly, increased sarcosine levels may boost cognition, as previously shown in patients with schizophrenia, in which oxidative damage and impaired glucose metabolism play key roles (*91*). In addition, decreased histidine metabolism and other decreased markers, such as homocysteine and S-adenosylhomocysteine (SAH) found in our treatment group, have been already shown to slow the cognitive ageing process appropriately downregulated (*92*). For instance, increased plasma homocysteine levels are a known risk factor for AD, whereas a low leucine and arginine diet yield beneficial cognitive effects (*93*).

Unexpectedly, CMAs rapidly lowered uric acid and associated metabolites levels. Uric acid stimulates inflammation either directly or by activating NLRP3 inflammasomes (*94*). Although the extent to which uric acid reduction contributed to the regression in cognitive impairment is unclear, it likely that it is linked to the improvement in the metabolic homeostasis. A good example is a recent clinical study showing increased urea metabolism in patients with AD (*95*). Accordingly, decreased taurine levels and urea metabolites are associated with a diminished risk of dementia (*96*). The majority of clinical study findings collectively agree with our results, showing significantly dysregulated baseline metabolites, which normalized with treatment.

Of note, to date, a few studies aimed to identify global changes in metabolites and metabolic pathways in AD (*15, 97, 98*). Among these, some studies highlighting that lipid dysfunction also plays an important role in the pathophysiology of AD (*99*). In terms of lipid metabolism, significant differences in the levels of some compounds have been observed in patients with AD. Despite some discrepant trends in cross-sectional studies examining the levels of lipids in AD patients (*100, 101*), the plasma levels of sphingolipids, sphingomyelins(*102, 103*), acylcarnitines (*104*)and phosphatidylcholines(PC) (*105–107*) exhibited statistically lower concentrations in patients with AD, even in the preclinical stages of the disease (*18*). In addition, a significant correlation among different lipid metabolites, tau and amyloid pathology, brain atrophy and cognitive decline was observed in a AD human study (*18*). An autopsy study of frontal cortex metabolites from patients with AD showed that impaired glycerophospholipid metabolism was involved in six central metabolic pathways reported to be altered in the disease (*108*). In brief, we observed significantly increased post-therapeutic levels of lipid metabolites, previously reported to decrease in patients with AD, including sphingomyelin, carnitine and carnitine-related by-products.

Despite insufficient clinical AD data concerning cholesterol metabolites and dicarboxylic acids (DCAs), we observed significantly lower levels of these metabolites after CMA treatment (*109*). Levels of pregnanediol, a metabolite of pregnenolone, and DCAs, end-products of β- or omega oxidation, which were observed as decreased in the present study, were previously reported to be lower in the urine of patients with AD (*110, 111*). Considering the neurotoxic role of bile acids, along with the oxidative properties of DCAs, the detection of decreased levels of bile acid metabolites and DCA products in the present study is therefore not surprising. Similarly, allopregnanolone has already been reported to result in deleterious effects on cognitive functions through gamma-aminobutyric acid (GABA) signalling (*112*). Also, increased bile acid levels have been reported in mild cognitive impairment (MCI) and AD. In contrast, bile acids strongly inhibited the cysteine catabolic pathway in the preclinical period, resulting in depletion of the free cysteine pool and reduction of antioxidant glutathione concentrations(*113*).

Although there has been no direct evidence relating plasma ascorbic acid (AA) levels to the pathogenesis of AD, our finding of decreased plasma levels after CMAs may be related to increased brain concentrations and central consumption of ascorbic acid in cognitively improved patients with AD. In this respect, and contrast to other metabolites, a direct transmission from the periphery to the brain suggested that AA might play a ‘nourishing’ role in the brain. A direct correlation between brain levels of AA and dementia was eventually confirmed by a recent study showing that a high CSF: plasma AA ratio was a marker of a “healthier” brain, better able to cope with the neurodegenerative process in AD (*114*).

However, despite these promising human data, plasma levels of several proteins in AD observed after CMA treatment in the present study were inconsistent with their established beneficial role under experimental conditions. For example, PSP, which decreased in the present study, is a novel neurotrophic factor exhibiting significant similarities to GDNF by exerting intense neurotrophic activity, specifically on central neurons (*115, 116*). Such discrepancies are not easily explained, even if the different results obtained in peripheral blood can be correlated with improved clinical status. A finding of decreased or increased post-therapeutic levels of these proteins in patients with AD may reflect a treatment-related alteration in their cellular processing in the central nervous system, resulting in variability in protein production and/or degradation in peripheral blood. In addition, controversial data have been reported regarding the concentrations of most of these proteins in patients with AD (*102, 117–120*). Also, due to the high heterogeneity of clinical cohorts and the restricted number of patients, the reproducibility of previous proteomics studies’ results is noticeably low.

Our therapeutic data fits well with recent ROSMAP transcriptomic data in AD patients, showing impaired glycolysis, urea cycle, and fatty acid metabolism that was considerably normalized after the CMA treatment. For instance, our transcriptomic analysis showed increased expressions of pyruvate dehydrogenase kinase 4 (PDK4), carnitine palmitoyltransferase (CPT1A) and hexokinase 2 (HK2). It decreased expressions of acyl-CoA dehydrogenases (ACADs), ATP synthase (ATP5MGL), and acetyl- and acyltransferases (GCNT7, MBOAT4, GALNT17) which have been already reported to link/associate with critical energetic deficiencies due to decreased glycolysis and lipid oxidation metabolism. Consistent with the above-mentioned expression levels, we observed increased metabolites in the cytosol, including carnitine, spermine, serine, alanine, glucose 6-phosphate, and acyl-CoA responsible for glycolytic and lipid metabolism-related energetic cascades. Also, our animal data confirmed the beneficial effects of our CMAs, showing the most substantial impacts on hyperemia, neuronal degeneration and necrosis in HFD animals.

A few limitations of the study need to be considered. First, the treatment effect was assessed by clinical evaluation and omics-analysis. Thus, our findings warrant a clinical trial with neuroimaging analysis to delineate the impact of CMAs on functional and structural brain alterations. Second, the link between systemic and CNS alterations and their relations to the AD pathology, i.e., amyloid and tau aggregation, has not been evaluated. Thus, further amyloid or tau-based neuroimaging combined with CSF evaluation should be pursued.

In summary, the human phase 2 clinical study supports the data from animal models and genome-scale metabolic modelling suggesting that oral administration of metabolic activators can improve the mitochondrial dysfunction in AD patients. The safety profile of metabolic activators in these patients was consistent with the results of our previous one-day calibration study and clinical trials, including only a single component of the CMAs (*20*). Our present study showed that CMAs was safe and well-tolerated in patients with AD, and no major safety concerns were identified. Importantly, CMAs improved cognition and serum markers in these patients after only 12 weeks of treatment. These findings suggest that targeting multiple pathways by metabolic activators is a potentially effective therapeutic strategy for AD.

## MATERIAL AND METHODS

### Transcriptomics data analysis of the brain in AD patients

The mRNA expression profiles of 629 AD and 704 control samples were obtained from the Religious Orders Study and Rush Memory Aging Project (ROSMAP) datasets (*26–28*). The data has been normalized by quantile scaling, TMM normalization, Pareto scaling, and then limma *removeBatchEffect* (*121*). DESeq2 (*122*) was used to identify the differentially expressed genes (DEGs) with uniform size factors. Ensembl Biomart (*123*) was used to mapping different gene accession IDs or symbols. Gene set enrichment (GSE) was performed by using piano (*124*). The top 5% DEGs by DESeq2 statistic were accepted for GSE analysis. To identify the significantly changed metabolites in AD, we performed the Reporter metabolite analysis based on the RAVEN Toolbox 2.0 *reporterMetabolites* function (*125*), which uses DEGs information through the network topology of the reference metabolic model. The *iBrain2845* (*29*) genome-scale metabolic model was used as the reference metabolic model. KEGG Pathway Mapper (*126*) was used to identify predicted changed pathways based on maps M01200, M01212, and M01230

### Animal study design

12-week old female Sprague-Dawley rats (weighing 320-380g) were kept at a controlled temperature of 22 ± 2°C and a controlled humidity of 50 ± 5% on a 12-hour light/dark cycle. Food and water were available ad libitum. The animal experiments are performed at Medical Experimental Research Center, Atatürk University, Erzurum, Turkey. All experiments for the treatment of the animals were approved by the Ethics Committee of Atatürk University, and were conducted following the National Institutes of Health Guide for Care and Use of Laboratory Animals.

After acclimation to laboratory conditions for 1 week, rats were randomly divided into two dietary regimens receiving either a Chow diet (CHOW) or a high-fat diet (HFD. Group 1 (n=4) were fed with only regular CD for 5 weeks; Group 2 (n=4) were fed with CD and treated with STZ. The remaining animals in Group 3 (n=4) were fed with HFD for 3 weeks and sacrificed to verify the model development. Groups 4-10 were fed with HFD for 5 weeks. After 3 weeks, the animals in Groups 5-10 were treated with STZ and administered with individual or combined metabolic activators, including L-serine, NAC, LCAT and NR for 2 weeks.

### Intracerebroventricular-streptozotocin (STZ) injection

Before surgical procedures, the rats were anesthetized by intraperitoneal administration of ketamine–xylazine (50 mg/kg ketamine and 5mg/kg xylazine) and placed individually in the stereotaxic instrument (Stoelting, Illinois, USA). Stereotaxic coordinates for injection were 0.8 mm posterior to the bregma, 1.5 mm lateral to the sagittal suture and 3.6 mm below the brain surface (*127*). Then, 10 microliters of STZ (3 mg/kg, Sigma-Aldrich, Darmstadt, Germany) were injected over 3 min with a Hamilton micro-syringe into the bilateral ventricle. The injection needles were left in place for an additional 2 min to allow diffusion.

### CMAs administration to animals

For carrying out the treatment experiments, the HFD rats were randomly separated into 9 (Groups 5-10) subgroups (n☐=☐4) and all treated with 3☐mg/kg STZ (10☐µL, icv). Group 5, treated only with STZ; Group 6, treated with STZ and serine (1000 mg/kg once daily by oral gavage); Group 7, treated with STZ and NAC (300 mg/kg once daily by oral gavage); Group 8, treated with STZ and LCAT (100 mg/kg once daily by oral gavage); Group 9, treated with STZ and NR (120 mg/kg once daily by oral gavage); Group 10, treated with STZ and serine (1000 mg/kg once daily by oral gavage), NAC (300 mg/kg once daily by oral gavage), LCAT (100 mg/kg/day) and NR (120 mg/kg once daily by oral gavage).

The body weight of the rats was recorded each week (Dataset S2). After 5 weeks, all animals were anesthetized with isoflurane and sacrificed. Blood samples were collected from the abdominal aorta and centrifuged at 8000☐rpm for 15☐min at 4☐°C for blood biochemistry analysis using automatic chemical analyser. The internal organs, including the heart, adipose tissues, liver, kidney, brain, muscle, intestine (duodenum, ileum, jejunum), pancreas, colon and stomach were immediately removed and then snap-frozen in liquid nitrogen and stored at ☐80°C.

Liver and brain tissue samples obtained due to the experimental procedure were fixed in 10% buffered formalin solution for 48 hours. Following the routine tissue procedure, the tissues were embedded in paraffin blocks and 4 µm thick sections were taken from each block. Preparations prepared for histopathological examination were stained with hematoxylin-eosin (HE) and examined with a light microscope (Olympus BX51, Germany). According to histopathological findings sections were evaluated by independent pathologist and scored as absent (-), very mild (+), mild (++), moderate (+++) and severe (++++).

### Immunohistochemical examination

Tissue sections taken on the adhesive (poly-L-Lysin) slides for immunoperoxidase examination were deparaffinized and dehydrated. After washing, the tissues with suppressed endogenous peroxidase activity in 3% H_2_O_2_ were boiled in antigen retrieval solution. To prevent nonspecific background staining in the sections, protein block compatible with all primary and secondary antibodies was dropped and incubated for 5 minutes. Caspase 3 (cat no:sc-56053 dilution ratio:1/100 US) for liver tissues and 8-OH-dG (Cat no: sc-66036 dilution ratio: 1/100 US) for brain tissues was used as the primary antibody. 3-3’ Diaminobenzidine (DAB) chromogen was used as chromogen, and according to their immunopositivity, sections were evaluated by independent pathologist and scored as absent (-), very mild (+), mild (++), moderate (+++) and severe (++++).

### Immunofluorescence examination

Tissue sections taken on the adhesive (poly-L-Lysin) slides for immunoperoxidase examination were deparaffinized and dehydrated. After washing, the tissues with suppressed endogenous peroxidase activity in 3% H_2_O_2_ were boiled in antigen retrieval solution. To prevent nonspecific background staining in the sections, protein block compatible with all primary and secondary antibodies was dropped and incubated for 5 min. For liver tissues, primary antibody 8-OHdG (Cat No: sc-66036 Dilution Ratio:1/100 US) was dropped and incubated at 37^°^C for 1 h. After washing, secondary FITC (Cat No: ab6717 Dilution Ratio:1/500 UK) was dropped and incubated at 37^°^C for 30 min. The other primary antibody H2A.X (Cat No:I 0856-1 Dilution Ratio:1/100 US) was dropped and incubated at 37^0^C for 1 h. After washing, secondary Texas Red (Cat No: sc-3917 Dilution Ratio:1/100 US) was dropped and incubated at 37^°^C for 30 min. DAPI (Cat No:D-1306 Dilution Ratio:1/200 US) was dripped onto the washed tissues and incubated in the dark for 5 min, then glycerine was sealed. For brain tissues, primary antibody Caspase 3 (Cat No:sc-56053 Dilution Ratio:1/100 US) was dropped and incubated for 1 h at 37^°^C. After washing, secondary FITC (Cat No: ab6717 Dilution Ratio:1/500 UK) was dropped and incubated at 37^°^C for 30 min. The other primary antibody H2A.X (Cat No: I 0856-1 Dilution Ratio:1/100 US) was dropped and incubated at 37^°^C for 1 h. After washing, secondary Texas Red (Cat No: sc-3917 Dilution Ratio:1/100 US) was dropped and incubated at 37^°^C for 30 min. DAPI (Cat No:D-1306 Dilution Ratio:1/200 US) was dripped onto the washed tissues and incubated in the dark for 5 min, then glycerine was sealed. Sections were examined under a fluorescence microscope (ZEISS Germany) by an independent pathologist and, according to their immunopositivity evaluated as absent (-), very mild (+), mild (++), moderate (+++) and severe (++++).

### Clinical Trial Design and Oversight

Patients for this randomized, double-blinded, placebo-controlled, phase 2 study were recruited at the Faculty of Medicine, Alanya Alaaddin Keykubat University, Antalya, Turkey and Faculty of Medicine, Istanbul Medipol University, Istanbul, Turkey. Written informed consent was obtained from all participants before the initiation of any trial-related procedures. The safety of the participants and the risk–benefit analysis was overseen by an independent external data-monitoring committee. The trial was conducted following Good Clinical Practice guidelines and the principles of the Declaration of Helsinki. The ethics committee approved the study of Istanbul Medipol University, Istanbul, Turkey, and retrospectively registered at https://clinicaltrials.gov/ with Clinical Trial ID: NCT04044131.

### Participants

Patients were enrolled in the trial if they were over 50 years of age with mild to moderate AD according to ADAS-cog (AD Assessment Scale-cognitive subscale; ADAS≥12) and the Clinical Dementia Rating Scale Sum of Boxes (CDR-SOB; CDR≤ . Patients who had a 2) history of stroke, severe brain trauma, toxic drug exposure were excluded. The main characteristics of the patients are summarized in Dataset S3. The inclusion, exclusion, and randomization criteria are described in detail in the Supplementary Appendix.

### Randomization, Interventions, and Follow-up

Patients were randomly assigned to receive CMAs or placebo (2:1). Patient information (patient number, date of birth, initials) was entered into the web-based randomization system, and the randomization codes were entered into the electronic case report form. All clinical staff were blinded to treatment, as were the participants.

Treatment started on the day of diagnosis. Both placebo and CMAs were provided in powdered form in identical plastic bottles containing a single dose to be dissolved in water and taken orally one dose in the morning after breakfast and one dose in the evening after dinner. Each dose of CMAs contained 3.73 g L-carnitine tartrate, 2.55 g N-acetylcysteine, 1 g nicotinamide riboside chloride, and 12.35 g serine. All patients received one dose during the first 28 days and received two doses until Day 84. All patients came for a follow-up visit on Day 84. Further information is provided in the Supplementary Appendix.

### Outcomes

The primary endpoint in the original protocol was to assess the clinical efficacy of CMAs in AD patients. For the primary purpose, the clinical differences in cognition of subjects receiving twelve-week treatment either with metabolic activators supplementation or placebo were determined. The primary analysis was on the difference in cognitive and daily living activity scores between the placebo and the treatment arms, which were assessed by Mini-Mental State Examination (MMSE), AD Assessment Scale-cognitive subscale (ADAS-Cog) and AD Cooperative Study - Activities of Daily Living (ADCS-ADL) in AD patients. The secondary aim in this study was to evaluate the safety and tolerability of CMAs. All protocol amendments were authorized and approved by the sponsor, the institutional review board or independent ethics committee, and the pertinent regulatory authorities.

Number and characteristics of adverse events, serious adverse events, and treatment discontinuation due to CMAs were reported from the beginning of the study to the end of the follow-up period as key safety endpoints. The changes in vital signs baseline values, and the status of treatment were recorded at Day 0 and 84. A complete list of the end points is provided in the Supplementary Appendix.

### Proteomics Analysis

Plasma levels of proteins were determined with the Olink panel (Olink Bioscience, Uppsala, Sweden). Briefly each sample was incubated with DNA-labelled antibody pairs (proximity probes). When an antibody pair binds to its corresponding antigens, the corresponding DNA tails form an amplicon by proximity extension, which can be quantified by high-throughput, real-time PCR. Probe solution (3 μl) was mixed with 1 μl of sample and incubated overnight at 4°C. Then 96 μl of extension solution containing extension enzyme and PCR reagents for the pre-amplification step was added, and the extension products were mixed with detection reagents and primers and loaded on the chip for qPCR analysis with the BioMark HD System (Fluidigm Corporation, South San Francisco, CA). To minimize inter- and intrarun variation, the data were normalized to both an internal control and an interplate control. Normalized data were expressed in arbitrary units (Normalized Protein eXpression, NPX) on a log2 scale and linearized with the formula 2NPX. A high NPX indicates a high protein concentration. The limit of detection, determined for each of the assays, was defined as three standard deviations above the negative control (background).

### Untargeted Metabolomics Analysis

Plasma samples were collected on Days 0 and 84 for nontargeted metabolite profiling by Metabolon (Durham, NC). The samples were prepared with an automated system (MicroLab STAR, Hamilton Company, Reno, NV). For quality control purposes, a recovery standard was added before the first step of the extraction. To remove protein and dissociated small molecules bound to protein or trapped in the precipitated protein matrix, and to recover chemically diverse metabolites, proteins were precipitated with methanol under vigorous shaking for 2 min (Glen Mills GenoGrinder 2000) and centrifuged. The resulting extract was divided into four fractions: one each for analysis by ultraperformance liquid chromatography– tandem mass spectroscopy (UPLC-MS/MS) with positive ion-mode electrospray ionization, UPLC-MS/MS with negative ion-mode electrospray ionization, and gas chromatography– mass spectrometry; one fraction was reserved as a backup.

### Determination of clinical variables informing response to CMAs administration

The patient groups with low and high levels of each clinical parameter were established based on the median score for that clinical parameter across all patients on Day 0. Patients scoring at or below the median were placed in the low group; patients scoring above the median were placed in the high group. ADAS-Cog scores were measured over different time points and statistical significance was tested between time points by using a paired t-test. Clinical parameters were deemed informative for the response to CMAs if exactly one group (low or high) exhibited more statistically significant changes in ADAS-Cog in the CMA group than in the placebo group.

### Statistical Analysis

Paired t-test was used to identify the differences in clinical parameters between time points and one-way ANOVA was used to find the shifts between CMA and placebo groups at each time point. For analysis of plasma metabolomics, we removed the metabolite profiles with more than 50% missing values across all samples. Metabolite changes between time points were analysed by paired t-test. Metabolite changes between CMA and placebo groups were analysed by one-way ANOVA. Missing values were removed in pairwise comparison. The p-values were adjusted by Benjamini & Hochberg method. Metabolites with a false-discovery rate of 5% were considered statistically significant. Two-sided Student’s t-test was used for statistical analyses of plasma parameters in animal model, statistical significance was considered p < 0.05.

For analysis of plasma proteomics, we removed the protein profiles with more than 50% missing values across all samples. Pair t-test was used to identify the changes between time points and one-way ANOVA was used to identify the changes between different groups. p<0.01 was considered statistically significant. Spearman correlation analysis was used to analyse the association between CMAs and clinical parameters or metabolomics or proteomics.

### Generation of Multi-Omics Network

Multi-omics network was generated based on the Spearman correlations and the significant associations (FDR < 0.05) are presented. The analyses were performed with SciPy package in Python 3.7. Centrality analysis on the network was performed using iGraph Python.

## Data Availability

All available data included in the supplementary datasets.

## ACKNOWLEDGMENTS

This work was financially supported by ScandiBio Therapeutics and Knut and Alice Wallenberg Foundation. The authors would like to thank the Metabolon Inc. (Durham, USA) for generation of metabolomics data, and ChromaDex Inc. (Irvine, CA, USA) for providing NR. AM and HY acknowledge support from the PoLiMeR Innovative Training Network (Marie Skłodowska-Curie Grant Agreement No. 812616) which has received funding from the European Union’s Horizon 2020 research and innovation programme.

## SUPPORTING INFORMATION

Supporting Information includes 8 Supplementary Datasets.

## Supplementary datasets

**Dataset S1.** Transcriptomic analysis of Alzheimer’s disease brain tissue samples in ROSMAP datasets.

**Dataset S2.** Study groups, body weight of animals and statistical analysis results of animal study

**Dataset S3.** Collection of samples of CMA and placebo groups and the measured values of clinical indicators before and after treatment.

**Dataset S4.** Statistical analysis of clinical indicators between different visits or groups.

**Dataset S5.** Plasma metabolomics data for each patient before and after treatment and statistical analysis of plasma metabolites between different visits or groups.

**Dataset S6.** The association between the plasma level of the four supplements serine, carnitine, cysteine and nicotinamide riboside with the plasma levels of other metabolites.

**Dataset S7.** Plasma proteomics data generated with the Olink cardiometabolic, inflammation, neurology and oncology panels for each patient before and after treatment and statistical analysis of plasma proteins between different visits or groups.

**Dataset S8**. Multi-Omics Network Data, including edges and nodes information. The network is presented in the iNetModels (http://inetmodels.com).

